# Does diet quality moderate the long-term effects of discrete but extreme PM_2.5_ exposure on respiratory symptoms? A study of the Hazelwood coalmine fire

**DOI:** 10.1101/2024.01.23.24301688

**Authors:** Thara Govindaraju, Martin Man, Alice J Owen, Matthew Carroll, Brigitte M. Borg, Catherine L Smith, Caroline X Gao, David Brown, David Poland, Shantelle Allgood, Jillian F Ikin, Michael J Abramson, Tracy A McCaffrey, Tyler J Lane

## Abstract

In 2014, a fire at an open cut coalmine in regional Victoria, Australia burned for 6 weeks. Residents of the nearby town of Morwell were exposed to smoke, which included high levels of fine particulate matter (PM_2.5_). We investigated whether the long-term effects of PM_2.5_ on respiratory health were moderated by diet quality. A cross-sectional analysis was conducted of data collected 8.5 years after the mine fire from 282 residents of Morwell and 166 residents from the nearby unexposed town of Sale. Primary outcomes were respiratory symptoms. Exposure was coalmine fire-related PM_2.5_ and diet quality was assessed with the Australian Eating Survey (AES). The moderating effect of diet quality on respiratory outcomes associated with PM_2.5_ was assessed using logistic regression models, adjusting for potential confounders. Diet quality was poor in this sample, with 60% in the lowest category of overall diet quality. Overall diet quality and fruit and vegetable quality significantly attenuated the association between PM_2.5_ and prevalence of chronic cough and phlegm. Sauce/condiment intake was associated with a greater effect of PM_2.5_ on COPD prevalence. No other moderating effects were significant. The moderating effects of overall diet quality and vegetable and fruit intake aligned with *a priori* hypothesis, suggesting potential protective benefits. Considering the poor-quality diet in this community, we would encourage better eating habits including improving the fruit and vegetable intake.

**Highlights:**

- Fire-related PM_2.5_ associated with higher prevalence of chronic cough and phlegm
- However, these effects were smaller with a higher-quality diet
- In line with *a priori* hypotheses, effect only observed among antioxidant-rich foods

## Introduction

The Hazelwood open cut brown coalmine in the Latrobe Valley of south-eastern Australia caught fire on 9^th^ February 2014. The nearby town of Morwell was covered in smoke and ash over a period of approximately six weeks [1]. The concentrations of fine particulate matter 2.5 µm or less in diameter (PM_2.5_) exceeded the average background PM_2.5_ concentrations of 6 µg/m^3^ and the modelled hourly mean reached as high as 3700 µg/m^3^ [2]. In response to community concerns around possible health impacts of the smoke on local residents, the Victorian State Government established the Hazelwood Health Study [1] (www.hazelwoodhealthstudy.org.au) to formally investigate long term health effects.

The impact of ambient PM_2.5_ on major health outcomes is well established, including increased risks for cardiovascular and respiratory diseases and all-cause mortality[3]. However, research is limited on long-term respiratory health consequences of extreme but medium duration PM_2.5_, defined as lasting weeks to months and typically from events such as prolonged bushfires, peat or coal mine fires. The historical analogues to the Hazelwood coalmine fire, including the 1952 Great Smog of London [4, 5] and the Donora smog of 1948 [6], suggest increased risk of developing acute respiratory conditions and death. In the absence of high-quality historical evidence, much of the evidence focuses on chronic exposures to ambient PM_2.5_, mostly originating from anthropogenic sources like transport, industry, and energy production [7]. Previously published analyses from the Hazelwood Health Study have found dose-response relationships between mine fire PM_2.5_ exposure in adults and increased odds of experiencing chronic cough, phlegm and current wheeze approximately 2.5 years after the event [8], and chronic cough and current wheeze up to 8.5 to 9 years after mine fire [9]. Between 3.5 to 4 years after the mine fire dose-response relationships were observed between PM_2.5_ and spirometry consistent with chronic obstructive pulmonary disease (COPD) in adult non-smokers [10].

There is growing evidence that nutrition is an important, modifiable risk factor in the development and management of asthma, COPD and lung cancer [11, 12]. Fruits and vegetables are of particular interest in respiratory health [13, 14] due to their nutrient profile including antioxidants, vitamins, minerals, fibre and phytochemicals such as flavonoids. A randomised trial found subjects who consumed a high fruit and vegetable diet for three months had a decreased risk of asthma exacerbations, compared to subjects who consumed a low fruit and vegetable diet [15]. Reduced risk of childhood wheezing and of asthma in adults and children with higher consumption of fruit and vegetables have also been shown in a meta-analysis [16].

Diet also moderates the harmful effects of chronic exposure to air pollutants such as PM_2.5_ [17], including on respiratory diseases [18]. The antioxidants, especially vitamin C, carotenoids and flavonoids found in fruits and vegetables, may prevent endogenous and exogenous oxidative damage caused by PM_2.5_ [19–22]. Antioxidant supplementation may also reduce the adverse effects of air pollution and inflammatory responses [23] and a protective effect of fruit and vegetables has been observed on lung function in children exposed to air pollutants [24]. However, research on the role of diet in moderating effects of PM_2.5_ (ambient or extreme) on respiratory health is limited.

Considering the gaps in the evidence, the current study investigated the potential moderating effects of diet on longer-term respiratory health outcomes in adults with or without exposure to medium­ duration high-intensity PM_2.5_, 8.5-9 years after the coalmine fire.

## Methods

The study is a cross-sectional analysis of follow-up survey data from the Hazelwood Health Study Adult Cohort [25]. Research questions and hypotheses were pre-registered on the Open Science Framework [26]. Notably, we highlighted both fruit and vegetable intake as the foods most likely to have a protective effect against the adverse health impacts of smoke exposure.

### Study population and design

The Hazelwood Health Study adult cohort comprised 4,056 participants from the Adult Survey which ran between May 2016 and February 2017. Participants had to have been residents of Morwell (exposed) or of the comparison town, Sale, located 64 km to the east (minimally or unexposed) and aged 18 years or older on 31 March 2014[25]. This analysis focuses on a follow-up survey conducted in late 2022, or 8.5-9 years after the coalmine fire. Living cohort members were eligible if they had a current email or phone number, had not refused further contact and were not randomly selected to participate in the concurrent Hazelwood Health Study Psychological Impacts Stream surveys [27], leaving 2,385 potential participants. Recruitment began in August 2022 with email and/or SMS invites and ended in December that same year.

### Respiratory assessment

Participants completed an online respiratory health survey using Redcap [28, 29] for data collection, which included a modified version of the self-report European Community Respiratory Health Survey (ECRHS) Ill Short Screening questionnaire [8]. The survey collected the presentation (“Yes” vs “No“) of respiratory symptoms including current wheeze, chest tightness, nocturnal and resting shortness of breath, current nasal allergies, as well as chronic respiratory symptoms (on most days for as much as three months a year) including chronic cough and chronic phlegm. The survey also measured self­ reported asthma and COPD, and date of diagnosis which was used to classify as pre- or post-mine fire.

### Mine fire-related PM_2.5_ exposure estimation

Individual exposures to coalmine fire-related PM_2.5_ were estimated using self-reported time-location diary data from the Adult Survey blended with modelled PM_2.5_concentrations [25]. The Commonwealth Scientific and Industrial Research Organisation (CSIRO) Oceans and Atmosphere estimated mine fire-related PM_2.5_ concentrations in the Latrobe Valley and surrounding areas through models incorporating variables such as wind direction, speed and temperature and the amount of PM_2.5_ released per unit mass of burning coal [2].

### Diet quality

Diet quality was assessed based on Australian Recommended Food Scores (ARFS) measured using the Australian Eating Survey Food Frequency Questionnaire (AES FFQ) [30]. The AES FFQ is a 120-item self­ administered questionnaire designed to collect information about dietary intake over the participants’ previous 6 months. It is semi-quantitative, providing a standard portion size for each food item determined using ’natural’ serving size (e.g., a slice, a cup), where possible. An individual response for each food, or food type, was required, with frequency options ranging from ’Never’ to ’4 or more times per day’, varying depending on the food, with some drink items up to ’7 or more glasses per day’. The survey was proprietary software made available through the University of Newcastle [31] and the Australian Food Composition Database [32] was used to generate individual median daily macronutrient intakes and percent energy derived from macronutrients.

ARFS were calculated using a subset of 70 AES FFQ questions, which comprised eight food group subscales -vegetables (20-item), fruit (12-item), meat protein (7-item), non-meat/vegetarian protein (6-item), grains (breads and cereals, 12-item), dairy (10-item), water (1-item) and sauces/condiments (2-item) [31] and the total score was calculated by summing the points for each item (ranges 0-73 points). Diet quality was categorised as “Needs work” (total score <33), “Getting there” (33-38), “Excellent” (39-46) or “Outstanding” (47+).

### Confounders

Demographic data were collected as part of the original Adult Survey and included age, sex, highest education (“secondary up to year 10,” “secondary years 11-12,” and Certificate/Diploma/tertiary degree” and smoking (smoking status and cigarette pack-years). Socioeconomic status was determined from the Index of Relative Socio-economic Advantage and Disadvantage (IRSAD), a socioeconomic score for residential areas from the 2016 Australian Census linked to participant’s residence at Statistical Area Level 2 [33].

### Statistical analysis

Simple descriptive statistics were used to summarise the cohort characteristics and outcomes in the exposed and non-exposed samples. The differences between those who were exposed to the mine fire (Morwell) and the comparison group (Sale) were assessed by Fisher’s Exact tests for categorical measures and Kruskal-Wallis tests for continuous measures, with significance set at p<0.05. Multiple logistic regression models were constructed to investigate associations between diet intake and respiratory health outcomes, as well as whether diet quality moderated the association between mine fire-related PM_2.5_ exposure and outcomes (via testing the interactions between diet and mine fire­ related PM_2.5_ exposure). In regression models, associations were measured per 10µg/m^3^ increment in daily mean PM_2.5_ exposure from the coalmine fire. Aside from water intake, which as a single-point scale was effectively binary, diet scores were scaled and mean-centred, meaning odds ratios reflected a one standard deviation increase in diet score.

Multiple imputations were used to account for the missing data for all regression models, with the number of imputations equivalent to the proportion of records with missing data (n=8 for 7.14% of missing data), and results were pooled according to Rubin’s rules [34]. In sensitivity analyses, we applied inverse probability weighting to account for attrition among those invited to participate in the survey. Both weighted and unweighted results were reported. Analyses were conducted in R [35] and SPSS (Ver 28, IBM Corp, Armonk, NY).

## Results

**Table 1** describes the characteristics of participants from Morwell and Sale. Morwell participants had lower socio-economic status and educational attainment. Eight years after the mine fire, the prevalence of all self-reported respiratory symptoms was still higher in Morwell than Sale (though the difference between groups in current nasal symptoms was non-significant). Prevalence of self­ reported asthma was comparable between the two groups, whereas the prevalence of self-reported COPD was slightly higher in Sale compared to Morwell.

**Table 1.** Participant characteristics across Morwell and Sale.

|  | Morwell (N=282) | Sale (N=166) | P-Value* |
| --- | --- | --- | --- |
| Age, years, | 58 [50, 65] | 57 [48, 66] | 0.309 |
| Sex - Female | 160 [57%] | 103 [62%] | 0.270 |
| Daily mean fire-related $PM_{2.5}$ exposure ($\mu g/m^3$) | 11 [7,18] | 0 [0,0] | <0.001 |
| Smoking status – Never | 134(48%) | 83(51%) | 0.128 |
| Former | 116(42%) | 73(45%) |  |
| Current | 29(10%) | 8(4.9%) |  |
| **Cigarette pack years (among current and former smokers)** | 12 [5, 23] | 20 [6, 27] | 0.162 |
| **Education** |  |  | 0.006 |
| Secondary up to year 10 | 67 [24%] | 20 [12%] |  |
| Secondary year 11-12 | 55 [20%] | 31 [19%] |  |
| Certificate/Diploma/Tertiary degree | 158 [56%] | 114 [69%] |  |
| **Residential socioeconomic score (IRSAD)** | 835 [796, 909] | 930 [844, 952] | <.001 |
| **Self-reported respiratory conditions: Pre-mine Fire** |  |  |  |
| Asthma | 85 [30%] | 38 [23%] | 0.100 |
| COPD | 6 [2.1%] | 2 [1.2%] | 0.710 |
| **Self-reported respiratory conditions & symptoms: 8 years post mine fire** |  |  |  |
| Asthma | 94 [33%] | 46 [28%] | 0.240 |
| COPD | 22 [7.9%] | 23 [14%] | 0.050 |
| Chronic cough | 112 [40%] | 35 [21%] | <0.001 |
| Chronic phlegm | 76 [27%] | 24 [14%] | 0.002 |
| Current wheeze | 142 [51%] | 60 [36%] | 0.003 |
| Chest tightness | 88 [31%] | 36 [22%] | 0.029 |
| Nocturnal shortness of breath | 70 [25%] | 22 [13%] | 0.002 |
| Shortness of breath while resting | 78 [28%] | 28 [17%] | 0.011 |
| Nasal allergy | 150 [54%] | 74 [45%] | 0.063 |

Values given as Median [Inter Quartile Range] or n (% of reporting population) unless otherwise specified. COPD – Chronic pulmonary obstructive disorder; IRSAD - Index of relative socio-economic advantage and disadvantage
\* Significance of difference through Kruskal Wallis tests for continuous variables and Fisher's Exact test for categorical variables with location as the grouping variable

### Diet quality of the cohort

The ARFS-rated diet quality was mostly similar in both groups except for higher-quality meat in Morwell and higher quality grain intake in Sale, as shown in Table 2. In both groups the percent total energy derived from macronutrients was slightly higher for fat and slightly lower for carbohydrate as compared to the acceptable macronutrient distribution range (15-25% of energy from protein, 20-35% from fat and 45-65% from carbohydrate) for Australia and New Zealand [36].

**Table 2.**
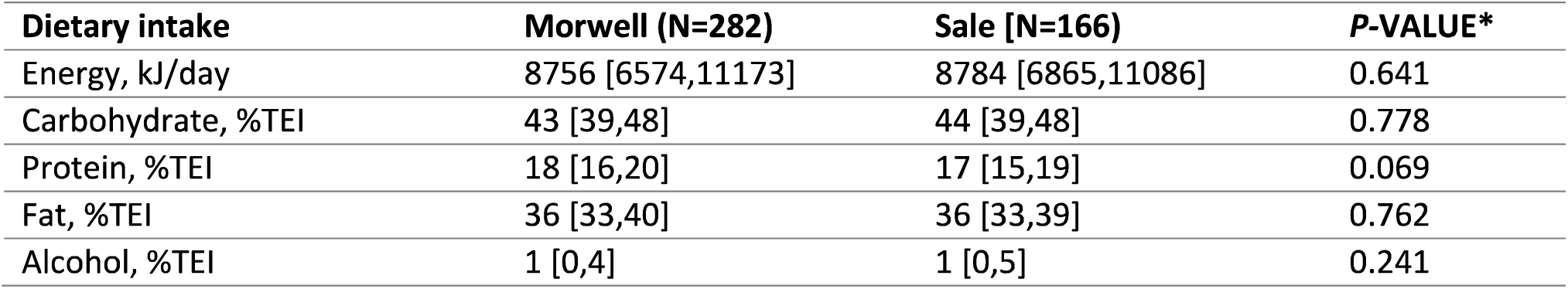
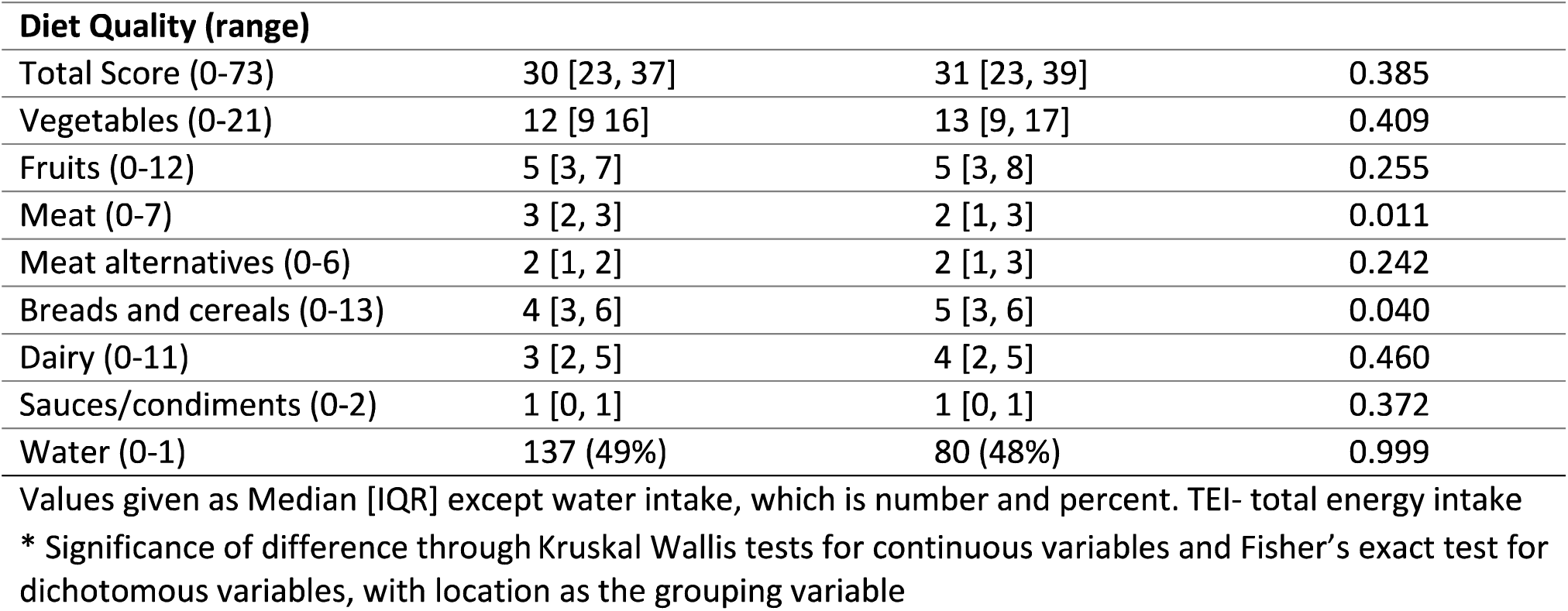
Dietary characteristics of participants across Morwell and Sale.

| Dietary intake | Morwell (N=282) | Sale [N=166] | P-VALUE* |
| --- | --- | --- | --- |
| Energy, kJ/day | 8756 [6574,11173] | 8784 [6865,11086] | 0.641 |
| Carbohydrate, %TEI | 43 [39,48] | 44 [39,48] | 0.778 |
| Protein, %TEI | 18 [16,20] | 17 [15,19] | 0.069 |
| Fat, %TEI | 36 [33,40] | 36 [33,39] | 0.762 |
| Alcohol, %TEI | 1 [0,4] | 1 [0,5] | 0.241 |

| **Diet Quality (range)** |  |  |  |
| --- | --- | --- | --- |
| Total Score (0-73) | 30 [23, 37] | 31 [23, 39] | 0.385 |
| Vegetables (0-21) | 12 [9, 16] | 13 [9, 17] | 0.409 |
| Fruits (0-12) | 5 [3, 7] | 5 [3, 8] | 0.255 |
| Meat (0-7) | 3 [2, 3] | 2 [1, 3] | 0.011 |
| Meat alternatives (0-6) | 2 [1, 2] | 2 [1, 3] | 0.242 |
| Breads and cereals (0-13) | 4 [3, 6] | 5 [3, 6] | 0.040 |
| Dairy (0-11) | 3 [2, 5] | 4 [2, 5] | 0.460 |
| Sauces/condiments (0-2) | 1 [0, 1] | 1 [0, 1] | 0.372 |
| Water (0-1) | 137 (49%) | 80 (48%) | 0.999 |

Values given as Median [IQR] except water intake, which is number and percent. TEI- total energy intake
\* Significance of difference through Kruskal Wallis tests for continuous variables and Fisher's exact test for dichotomous variables, with location as the grouping variable

Diet quality for most of the participants (60.1%) was in the range of “needing work” (<33), with very few (5.1%) in the “outstanding” range (47+), as shown in **Figure 1**.

**Figure 1.**
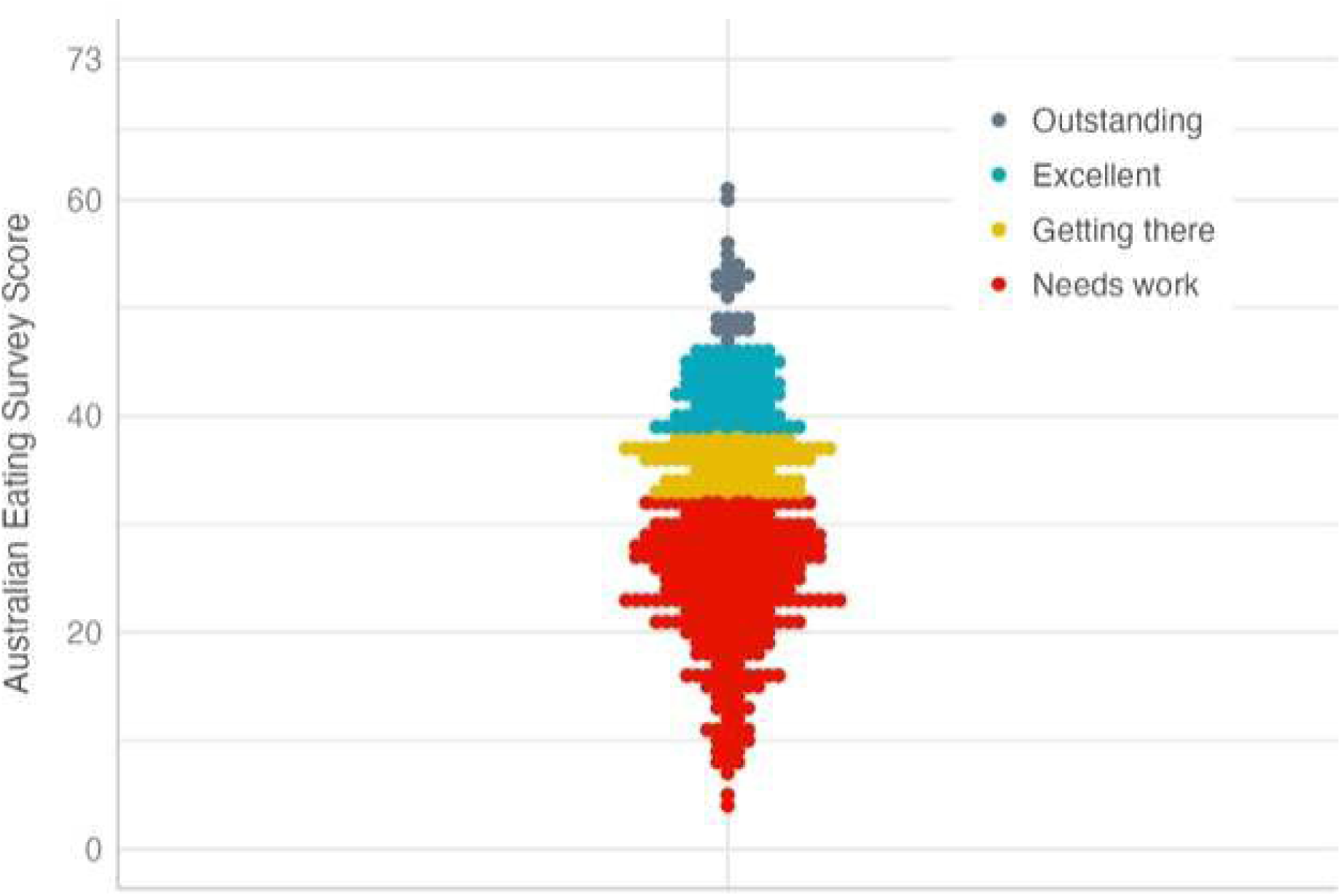
ARFS diet quality scores based on the Australian Eating Survey results for the whole cohort.

### Independent effects of fire-related PM_2.5_ and diet quality on respiratory health outcomes

We analysed the effects of fire-related PM_2.5_ and diet quality scores in separate models, while adjusting for confounders. In other words, the PM_2.5_ model did not adjust for diet quality and vice versa. Results are summarised in **Figure 2**.

**Fig 2.**
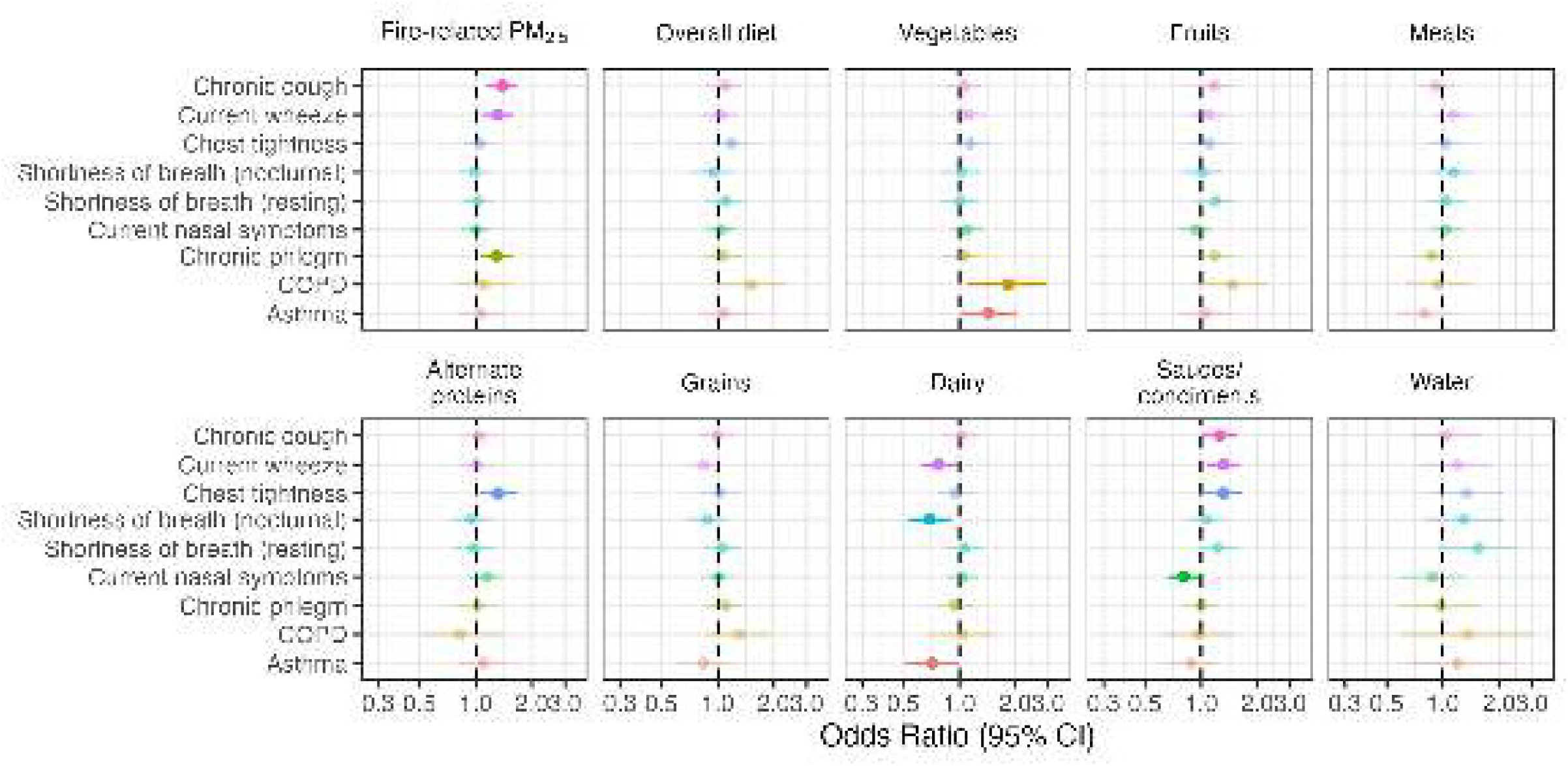
Forest plot showing the association between 10µg/m^3^ increases in fire-related PM_2.5_ and a standard deviation increase in diet quality score with the of reporting respiratory symptoms. Each row of ORs were estimated from an imputed multivariable logistic regression controlling for age, sex, smoking, IRSAD, and pre-fire asthma or COPD.

Fire-related PM_2.5_ was associated with higher prevalence of self-reported chronic cough, current wheeze, and chronic phlegm, after adjustment for confounders. Sauce/condiment intake was associated with higher prevalence of chronic cough, current wheeze, chest tightness, but reduced prevalence of current nasal symptoms. Dairy intake was associated with reduced prevalence of current wheeze, nocturnal shortness of breath, and asthma, while vegetable intake was associated with higher prevalence of COPD and asthma, and alternate proteins with higher prevalence of chest tightness.

### Moderating effects of diet quality on the association between PM_2.5_ and respiratory health outcomes

The respiratory symptoms that was most frequently moderated by the fire-related PM_2.5_ and diet interaction were chronic cough and chronic phlegm. Overall diet, and also vegetable and fruit quality, were all associated with reduced prevalence of chronic cough in association with fire-related PM_2.5_. Interestingly, in these models (Figure 3), the independent association between diet indicators and respiratory symptoms differed from the models in Figure 2, which did not adjust for fire-related PM_2.5_. In each case, the diet indicator predicted *higher* prevalence of chronic cough or chronic phlegm, although the effect was not always statistically significant. To understand these interactions better, we conducted further analyses between PM_2.5_and diet adjusting for confounders excluding smoking (Supplementary Figure 1). While no significant associations were found, point estimates were elevated for overall diet and vegetable and fruit quality, as well as meat quality. The implications will be discussed.

**Fig 3.**
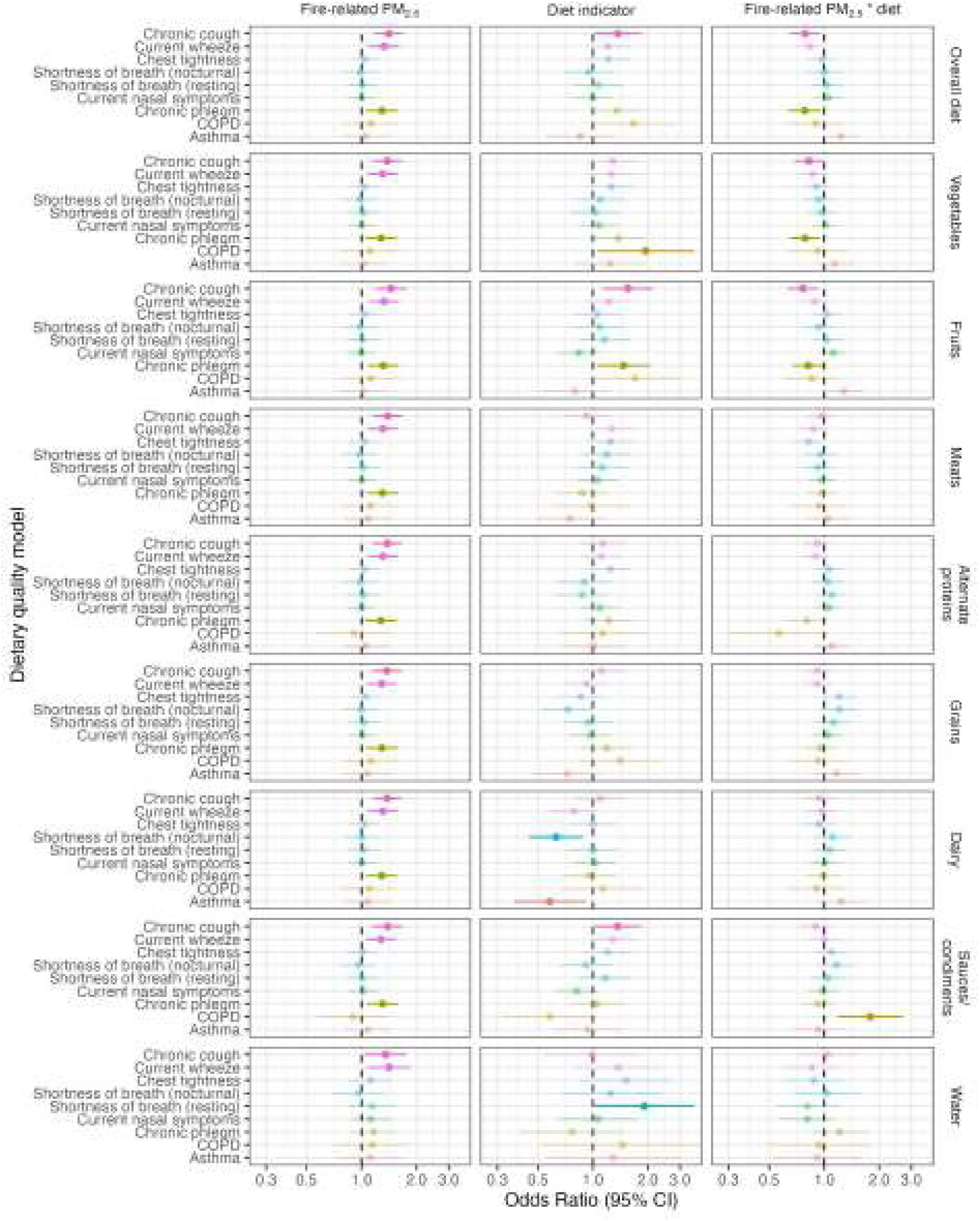
Forest plot depicting the interactions between 10µg/m^3^ increases in mine fire-related PM_2.5_ exposure and respiratory health outcomes, moderated by a standard deviation increase in diet quality score. Each row of ORs were estimated from an imputed multivariable logistic regression fitting mine-fire PM_2.5_ exposure, diet quality score and interaction between PM_2.5_ exposure and diet quality score controlling for age, sex, smoking and IRSAD score.

The only other significant interaction effect was sauces/condiments (tomato sauce and vegemite) intake, which significantly *increased* the prevalence of COPD in association with fire-related PM_2.5_.

These analyses are summarised in **Figure 3**.

**Supplementary Tables 1 and 2** provide detailed regression model outputs. We also conducted weighted analyses using inverse probability weighting which found fewer significant associations, although these were generally similar in direction **(Supplementary Figure 2).**

## Discussion

We hypothesised *a priori* that diet quality- and more specifically, fruit and vegetable intake - would offer protection against respiratory symptoms associated with smoke exposure from the coalmine fire [37]. Our findings generally supported this hypothesis; overall diet and fruit and vegetable intakes were associated with an attenuation of the effect of fire-related PM_2.5_ on the prevalence of chronic cough and phlegm. Fruit consumption has been suggested to have protective effects on respiratory health outcomes including inverse associations with cough and phlegm [38] and 25-year incidence of chronic nonspecific lung diseases characterised by chronic cough, phlegm, shortness of breath and wheezing[39]. While evidence strongly suggests fruit intake reduces the risk of asthma [40], we found no evidence of an effect. However, neither this study nor previous Hazelwood Health Study analyses [8] have found evidence that the mine fire exposures increased the prevalence of asthma; in other words, there may not be a harm to protect against.

There remains an apparent contradiction in our findings, with overall diet and fruit and vegetable quality seeming both to protect against the effect of fire-related PM_2.5_, while simultaneously increasing the prevalence of chronic cough and phlegm. As diet quality was measured years after the coalmine fire, reverse causality may be a factor; those most affected by the smoke - and who developed respiratory symptoms as a result - may have taken the biggest steps to improve their health, such as by eating better. Our analyses also indicated that while not significant, diet quality seemed to increase with exposure to fire-related smoke, after adjusting for all plausible confounders, barring tobacco use, as a change in diet could have been collinear with a change in smoking behaviours. The slight increase in diet quality with exposure could have been catalysed by health messaging through health care providers as a result of increased hospital visits or improved awareness through health promotion campaigns concerning diet and lifestyle, primarily organised as part of engaging with the community post-fire.

Several factors need consideration while interpreting the results of this study. The ARFS counts all vegetables equally, making no distinction on important characteristics such as antioxidant capacity that may influence protective effects [41]. For instance, while higher intake of fruit and vegetables in general is linked to reduced mortality, consumption of starchy vegetables like potatoes, which account for a quarter of vegetables consumed in Australia [42], is not [43]. Furthermore, while the epidemiological evidence for protective effects of fruits and vegetables is substantial, findings from randomised controlled trials have been mixed [44, 45].

Dairy quality was negatively associated with asthma and current wheeze independently but did not remain significant in the interaction model. The general perception of dairy worsening asthma and causing mucus, could have resulted in avoidance of dairy products and influenced the findings. The moderating effect of sauce/condiment intake on the PM_2.5_-COPD, which indicated increasing adverse effects of PM_2.5_, might have been a Type 1 Error, made more likely by the small numbers.

Most respondents were rated in the lowest diet quality category, “needs work.” Although similar to Australians generally [46], limited variability in ARFS score restricted our ability to identify moderating effects of diet quality. While overall diet quality of regional Australians was outside the scope of our study, this finding suggested there may be opportunities to improve general health in communities like Morwell and Sale through improvement of diet. Diet quality has multifactorial influences including health conditions and nutrition awareness, and socioeconomic factors, which are particularly relevant considering Morwell is in the 3^rd^ percentile nationwide and l5^1^ percentile in Victoria on disadvantage based on IRSAD scores[33]. The mechanisms by which a community may experience poor dietary health associated with mine fire exposure may include food scarcity, or lack of awareness regarding healthy eating and wider food environments structured on socio-economic status [47]. This could be exacerbated by limited access to health services due to living in a regional area.

To our knowledge, this is the first study to investigate moderating effects of diet quality on longer­ term respiratory health outcomes when exposed to extreme but medium-duration PM_2.5_. This topic is of growing importance as climate change is increasing the frequency and intensity of such events, such as the 2019/2020 Australian Black Summer bushfire season, the 2020 Western United States wildfire season, and the 2023 Canadian, Mediterranean and Hawaiian wildfires.

### Strengths and limitations

This research has substantial strengths, including the use of a validated diet survey, modelled smoke/PM_2.5_ exposure data, and an established cohort.

However, there are several limitations. PM_2.5_ exposure was based on time-location diaries collected 2.5 years after the mine fire and were thus subject to recall bias. Also, they did not account for protective behaviours such as wearing masks or staying indoors. Causality cannot be inferred owing to the nature of cross-sectional analysis. As a result of multiple comparisons, the significant associations must be interpreted cautiously and within the context of the current evidence.

## Conclusions

We found evidence that higher overall diet quality may protect or mitigate against the long-term effects of smoke exposure from a coalmine fire. Vegetable and fruit intake stood out, in line with our *a priori* hypotheses. Nevertheless, further confirmation from larger studies is needed to validate the findings. In the interim, larger studies could help generate greater certainty about the protective effects of diet quality against the harms of air pollution. In addition, there are broader policy implications to improving the quality of diet in areas that face numerous socioeconomic and health challenges, especially with the growing impacts of climate change.

## Data Availability

Hazelwood Health Study data are confidential and cannot be publicly shared. Analytical code has been archived on a public repository

https://bridges.monash.edu/articles/online_resource/Does_diet_quality_moderate_the_long-term_effects_of_discrete_but_extreme_PM2_5_exposure_on_respiratory_symptoms_A_study_of_the_Hazelwood_coalmine_fire_-_Analytical_code/23607288

**Statements and Declarations**

## Acknowledgements

We thank the Latrobe Valley and Gippsland communities for their support and participation in the Hazelwood Health Study.

## Ethics

Monash University Human Research Ethics Committee approved this study as part of the Hazelwood Adult Survey & Health Record Linkage Study (Project ID: 25680; previously CF15/872 - 2015000389 and 6066)

## Availability of data and material

Hazelwood Health Study data are confidential and cannot be publicly shared. Analytical code has been archived on a public repository [48].

## Supplements

### Supplementary tables

**Table S1.** Regression models for diet quality and PM_2.5_ across respiratory health outcomes.

|  | Chronic cough |  | Current wheeze |  | Chest tightness |  | Shortness of breath (nocturnal) |  | Shortness of breath (resting) |  | Current nasal symptoms |  | Chronic phlegm |  | Asthma |  | COPD |  |
| --- | --- | --- | --- | --- | --- | --- | --- | --- | --- | --- | --- | --- | --- | --- | --- | --- | --- | --- |
| Diet quality (ARFS) | OR (95%CI) | P-VALUE | OR (95%CI) | P-VALUE | OR (95%CI) | P-VALUE | OR (95%CI) | P-VALUE | OR (95%CI) | P-VALUE | OR (95%CI) | P-VALUE | OR (95%CI) | P-VALUE | OR (95%CI) | P-VALUE | OR (95%CI) | P-VALUE |
| Total Score | 1.08 (0.78-1.50) | 0.624 | 1.01 (0.74-1.39) | 0.94 | 1.27 (0.90-1.78) | 0.178 | 1.03 (0.71-1.48) | 0.882 | 1.16 (0.81-1.67) | 0.422 | 1.07 (0.79-1.44) | 0.673 | 1.04 (0.73-1.49) | 0.828 | 1.02 (0.62-1.68) | 0.945 | 1.64 (0.87-3.09) | 0.123 |
| Vegetables | 1.04 (0.75-1.43) | 0.812 | 1.08 (0.79-1.47) | 0.646 | 1.18 (0.84-1.65) | 0.345 | 1.07 (0.75-1.54) | 0.706 | 1.06 (0.74-1.51) | 0.763 | 1.12 (0.83-1.50) | 0.457 | 1.00 (0.70-1.42) | 0.997 | 1.44 (0.86-2.42) | 0.169 | 2.06 (1.01-4.18) | 0.047 |
| Fruits | 1.11 (0.80-1.55) | 0.526 | 1.03 (0.75-1.41) | 0.876 | 1.12 (0.80-1.57) | 0.516 | 1.02 (0.71-1.47) | 0.9 | 1.21 (0.84-1.74) | 0.303 | 0.94 (0.70-1.27) | 0.697 | 1.18 (0.82-1.69) | 0.382 | 1.06 (0.65-1.74) | 0.812 | 1.39 (0.75-2.58) | 0.29 |
| Meats | 0.93 (0.68-1.26) | 0.621 | 1.13 (0.83-1.52) | 0.437 | 1.06 (0.77-1.47) | 0.708 | 1.17 (0.82-1.65) | 0.385 | 1.04 (0.74-1.46) | 0.829 | 1.11 (0.84-1.48) | 0.454 | 0.86 (0.61-1.22) | 0.403 | 0.86 (0.53-1.40) | 0.538 | 1.10 (0.62-1.94) | 0.742 |
| Alternate proteins | 0.97 (0.68-1.39) | 0.887 | 0.97 (0.70-1.34) | 0.844 | 1.39 (1.00-1.93) | 0.052 | 1.04 (0.72-1.50) | 0.825 | 0.99 (0.68-1.44) | 0.949 | 1.22 (0.88-1.68) | 0.227 | 0.97 (0.65-1.45) | 0.884 | 0.92 (0.54-1.60) | 0.78 | 0.78 (0.36-1.68) | 0.518 |
| Grains | 1.10 (0.81-1.51) | 0.537 | 0.83 (0.60-1.13) | 0.233 | 1.10 (0.80-1.52) | 0.56 | 0.94 (0.66-1.33) | 0.718 | 1.08 (0.77-1.52) | 0.668 | 0.99 (0.74-1.33) | 0.969 | 1.13 (0.80-1.59) | 0.497 | 0.77 (0.47-1.24) | 0.282 | 1.22 (0.68-2.16) | 0.506 |
| Dairy | 1.06 (0.77-1.46) | 0.718 | 0.85 (0.63-1.16) | 0.31 | 1.01 (0.73-1.41) | 0.943 | 0.80 (0.55-1.14) | 0.215 | 1.10 (0.77-1.56) | 0.61 | 1.04 (0.77-1.39) | 0.815 | 0.97 (0.68-1.39) | 0.882 | 0.65 (0.39-1.09) | 0.101 | 1.06 (0.61-1.85) | 0.841 |
| Sauces/Condiments | 1.22 (0.89-1.67) | 0.207 | 1.23 (0.91-1.66) | 0.187 | 1.31 (0.95-1.80) | 0.099 | 1.04 (0.74-1.46) | 0.825 | 1.21 (0.86-1.70) | 0.268 | 0.78 (0.59-1.05) | 0.098 | 0.97 (0.69-1.36) | 0.868 | 0.85 (0.53-1.38) | 0.517 | 1.11 (0.62-1.99) | 0.719 |
| Water | 1.04 (0.67-1.63) | 0.848 | 1.19 (0.78-1.82) | 0.418 | 1.34 (0.85-2.11) | 0.207 | 1.29 (0.79-2.10) | 0.305 | 1.54 (0.95-2.49) | 0.077 | 0.88 (0.59-1.31) | 0.521 | 0.97 (0.59-1.57) | 0.891 | 1.19 (0.62-2.29) | 0.598 | 1.36 (0.61-3.01) | 0.448 |
| **Exposure to particulate matter** |  |  |  |  |  |  |  |  |  |  |  |  |  |  |  |  |  |  |
| PM2.5 | 1.38 (1.15-1.66) | **0.001** | 1.30 (1.08-1.57) | **0.006** | 1.04 (0.86-1.26) | 0.658 | 0.98 (0.79-1.22) | 0.884 | 1.02 (0.83-1.25) | 0.845 | 1.01 (0.84-1.20) | 0.954 | 1.29 (1.06-1.57) | **0.013** | 1.10 (0.78-1.56) | 0.581 | 1.06 (0.80-1.42) | 0.679 |

Notes- OR-odds ratio /ARFS-Australian Recommended Food Score/COPD- Chronic obstructive pulmonary disease; adjusted for age, sex, IRSAD and smoking /\*significant at 0.05 levels

**Table S2.**
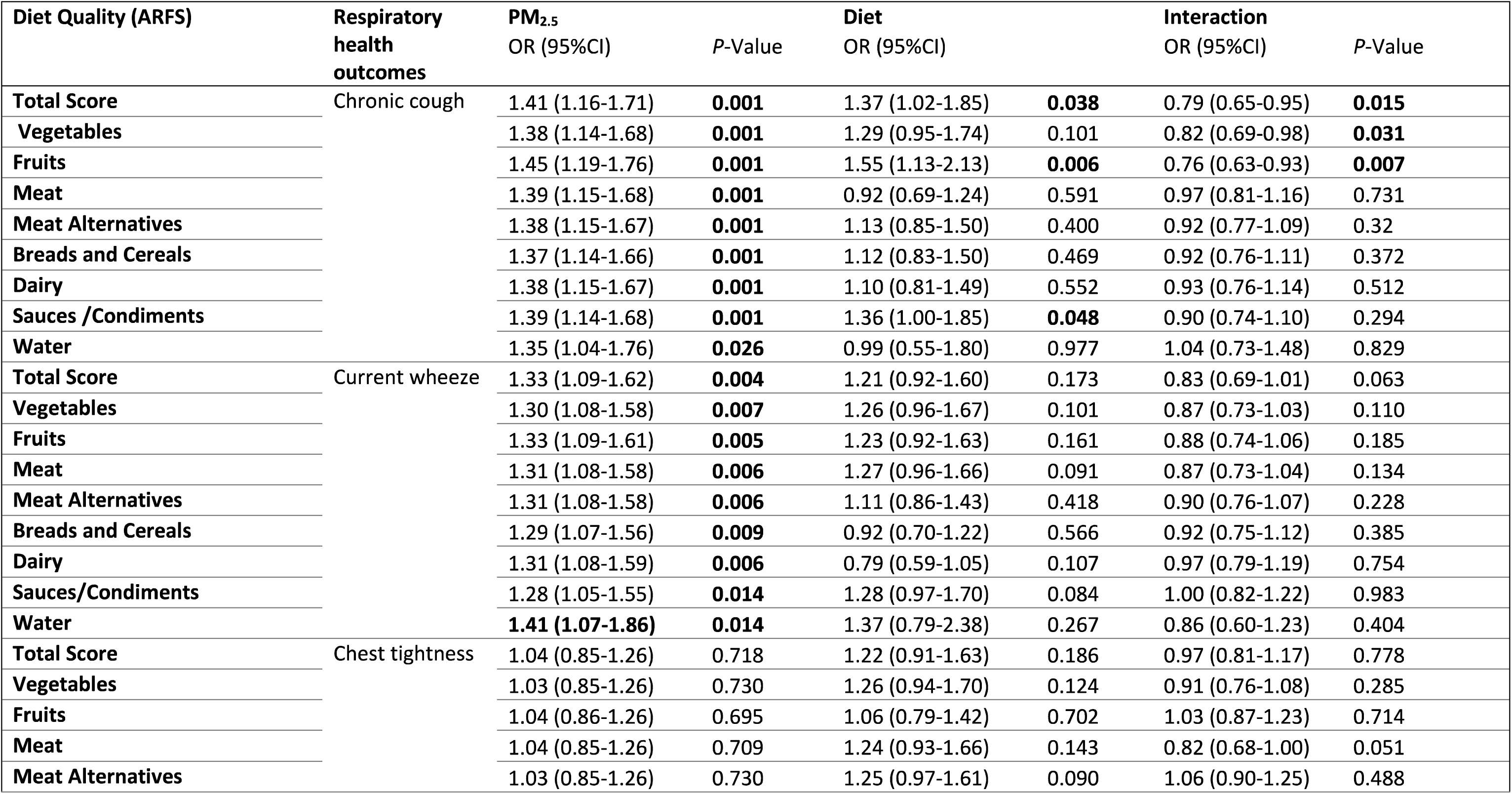
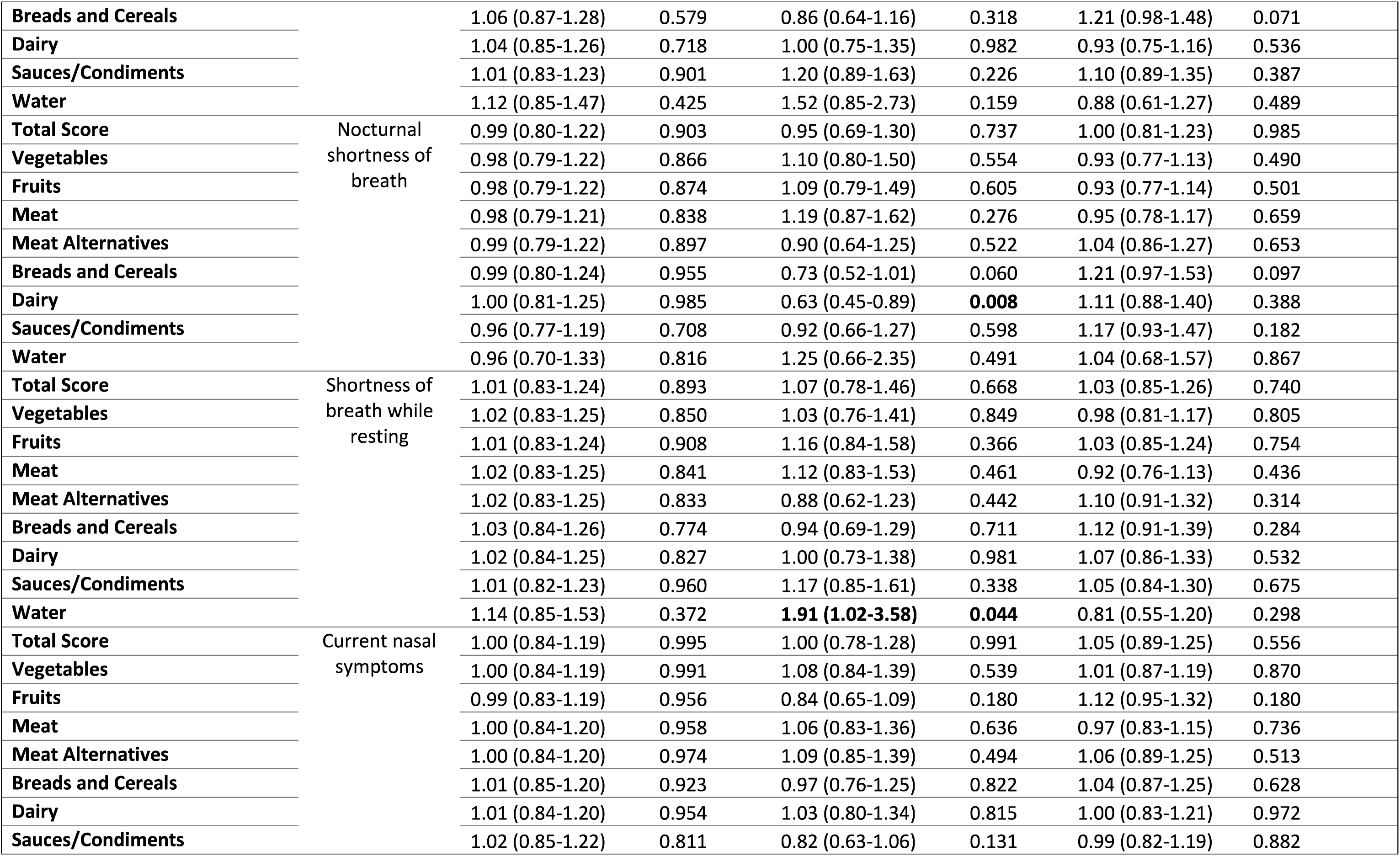
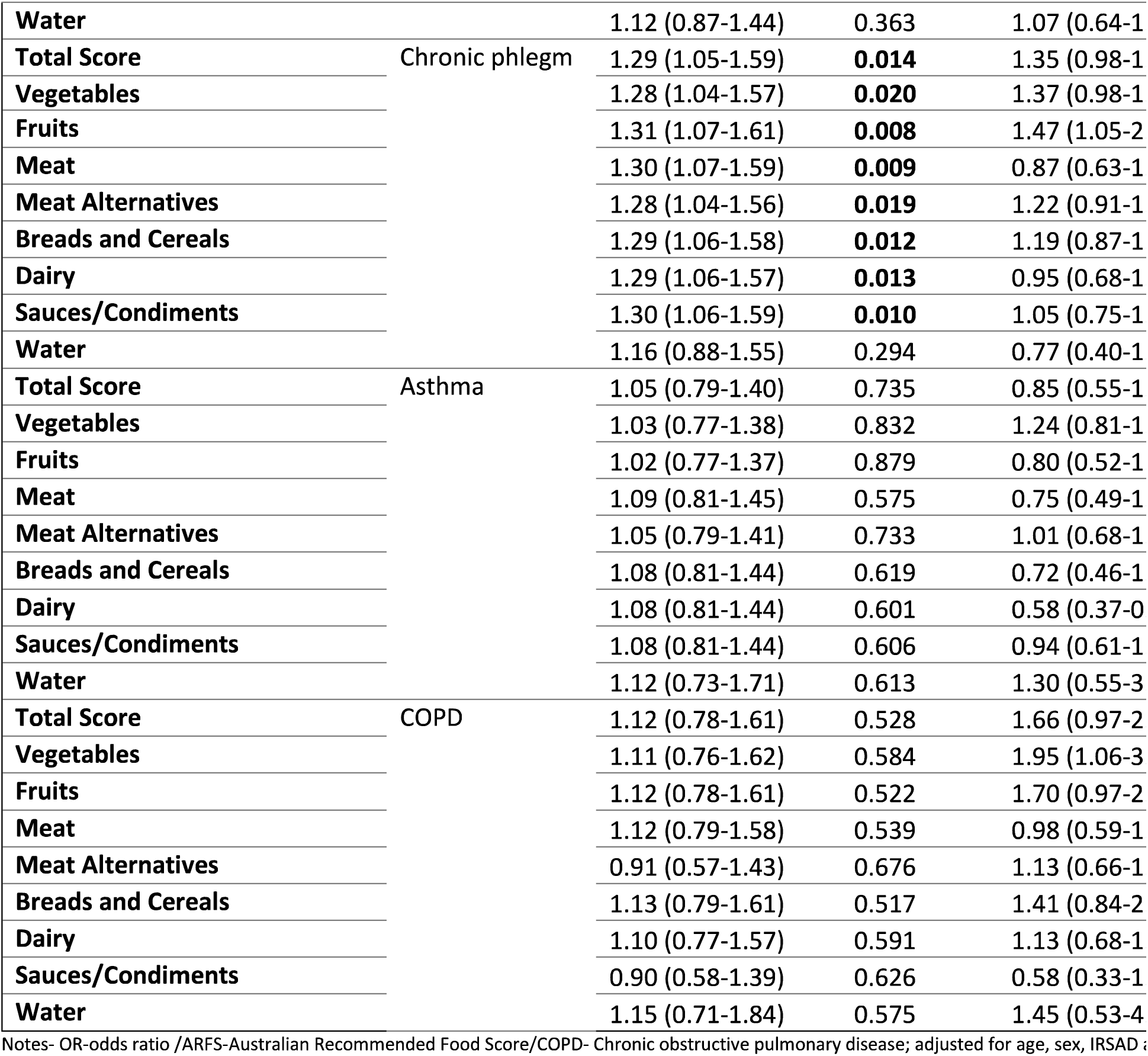
Regression models exploring the moderating role of diet quality between PM_2.5_ levels and respiratory health outcomes.

### Supplementary figures

**Figure S1.**
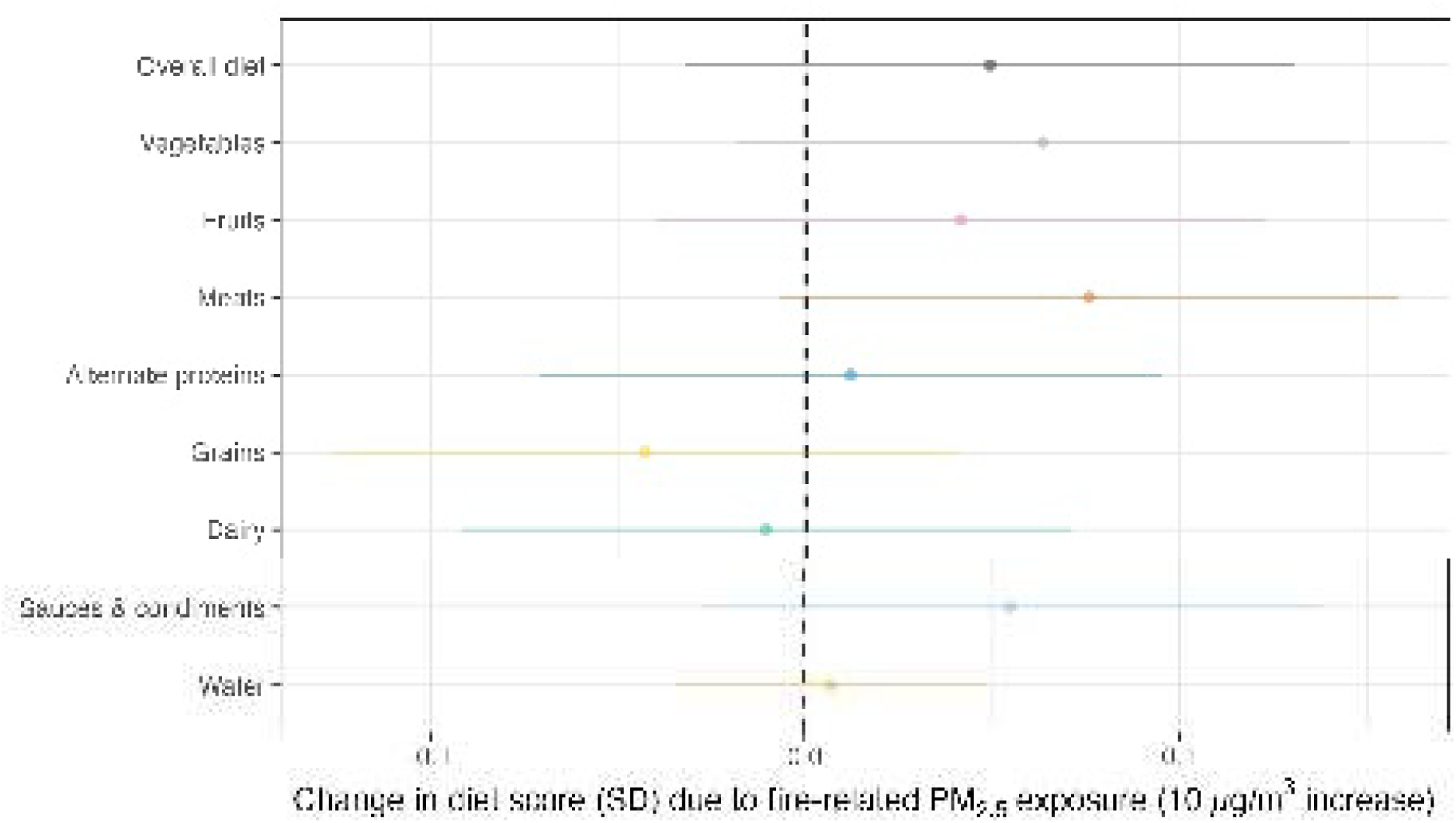
Change in Diet score (SD) due to fire-related PM_2.5_ exposure (10µg/m^3^ increase), adjusted for sex, age, educational attainment and IRSAD.

**Figure S2.**
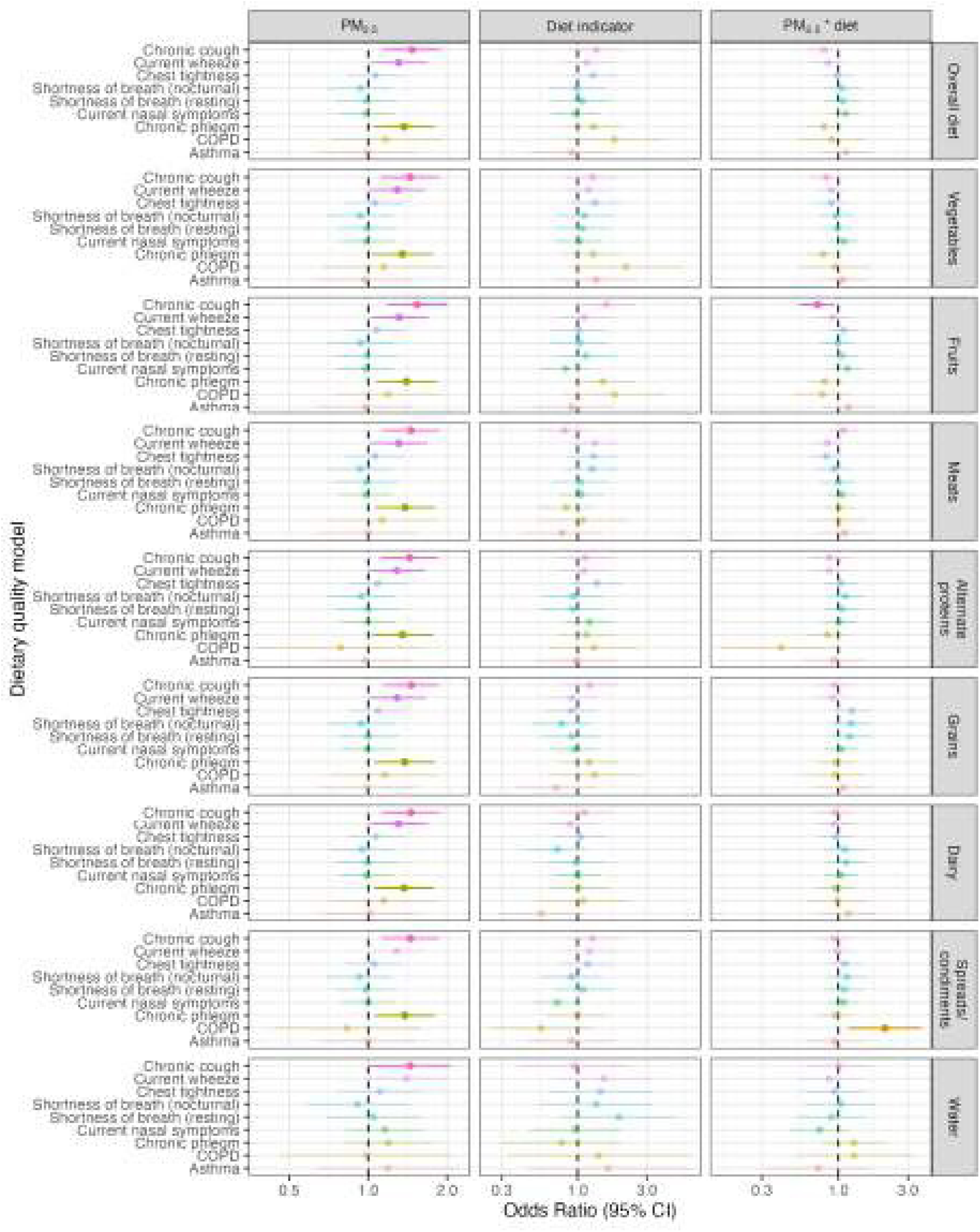
Forest Plot depicting the interaction coefficients for the association between 10µg/m^3^ of PM_2.5_ exposure from mine fires and respiratory health outcomes, moderated by diet quality; weighted analyses.

